# Smartphone ownership and use among pregnant women with HIV in South Africa

**DOI:** 10.1101/2022.09.29.22280417

**Authors:** Sandisiwe Noholoza, Tamsin K. Phillips, Sindiswa Madwayi, Megan Mrubata, Carol S. Camlin, Landon Myer, Kate Clouse

**Affiliations:** Division of Epidemiology and Biostatistics, School of Public Health and Family Medicine, University of Cape Town, Cape Town, South Africa; University of California, San Francisco, Department of Obstetrics, Gynaecology & Reproductive Sciences, San Francisco, CA, USA; Vanderbilt University School of Nursing, Vanderbilt University, Nashville, TN, USA; Vanderbilt Institute for Global Health, Vanderbilt University Medical Center, Nashville, TN, USA

**Keywords:** mHealth, smartphone, ownership, HIV, South Africa

## Abstract

**Background:** Mobile health (mHealth) initiatives are increasingly common in low-resource settings, but the appropriateness of smartphone interventions is uncertain. To inform future mHealth interventions, we describe smartphone ownership, preferences and usage patterns among women living with HIV (WLHIV) in Gugulethu, South Africa.

**Methods:** We screened pregnant WLHIV from December 2019 - February 2021 for the CareConekta trial. We describe sociodemographic characteristics and mobile phone ownership of all women screened (n=639), and smartphone use patterns among those enrolled in the trial (n=193).

**Results:** 91% owned a mobile phone; 87% of those owned smartphones. Among those with smartphones, 92% used Android operating system version 5.0 or above, 98% of phones had GPS and 96% charged their phones <twice/day.

Among 193 women enrolled, 99% owned the smartphone themselves; 14% shared their smartphone with someone but 96% of these possessed the phone most of the day. Median duration of smartphone ownership and current phone number was 12 [IQR:5-24] and 25 [IQR: 12-60] months, respectively. Participants reported a median of two [IQR: 1-2] phone numbers in the preceding two years.

Receiving (100%) and making (99%) phone calls were the most common smartphone uses; least used were GPS (55%) and email (47%). WhatsApp (94%) was the favourite app reported.

**Conclusion:** Smartphone ownership is very common in this low-resource, peri-urban setting. Phone sharing was uncommon, nearly all used the Android system and phones retained sufficient battery life. These results are encouraging to the development of mHealth interventions. Existing messaging platforms – particularly WhatsApp – are exceedingly popular and could be leveraged for interventions.

## Introduction

Mobile health (mHealth) interventions for the improvement of public health are increasing in popularity globally. The proliferation of mobile phone ownership in the past two decades, especially in developing countries, has been a massive catalyst for mHealth, with an estimated 7.26 billion of the world’s population owning mobile phones in 2022.^1-3^ mHealth interventions range from simple text message interventions to multi-interface mobile applications (apps) that can be used to disseminate health information, facilitate treatment adherence, facilitate patient engagement and retention, and collect patient data.^4-8^

In 2021, 7.7 million people in South Africa were living with HIV, the largest HIV epidemic in the world.^9^ Although great strides have been made in the reduction of HIV incidence, vertical HIV transmission prevention, and early antiretroviral therapy (ART) initiation, there remain large gaps in the HIV cascade, particularly among pregnant and postpartum women living with HIV (WLHIV), who are at increased risk of dropping out of HIV care.^10-11^ mHealth interventions seek to address these gaps through information, adherence support, stigma mitigation, and engagement in HIV care through sustained communication with the health facility.^2,12-14^

Text messaging is widely available at low cost, even in the most basic mobile phone types. Text messaging interventions have been used to improve engagement in HIV care and adherence in developing countries, including South Africa, through clinic appointment and medication reminders, health education messages, and case management.^15-20^ Smartphone interventions, including the use of social media apps, expand the reach of mHealth, particularly to young people who remain at high risk of HIV infection and poor HIV treatment outcomes.^21,22^

Kruse et al., noted that barriers to mHealth implementation fall under three categories: lack of infrastructure, lack of equipment, and the technology gap.^23^ To make informed evidence-based decisions in the design and implementation of mHealth interventions, it is crucial to understand mobile phone ownership and use patterns in the intended target population within the context of these barriers. The rapid acceleration of technology impacts the access, feasibility, efficacy and sustainability of current and future mHealth interventions. Therefore, the most current data on mobile phone/smartphone ownership patterns is necessary to improve current mHealth interventions and to inform future mHealth interventions that target various steps in the HIV cascade.

This research aims to describe smartphone ownership, preferences, and usage patterns among WLHIV in South Africa in order to inform future interventions. Given the known successes of simpler mHealth interventions, more detailed and current research is needed to establish the appropriateness and feasibility of integrating new mHealth modalities (novel apps and social media apps) in the South African context. Mitigating this gap in knowledge within the context of each target population has the potential to inspire the design of more targeted mHealth interventions, with potential for greater reach, uptake and long-term success.

## Methods

### Study design and setting

In this cross-sectional sub-study, we used data from the CareConekta randomized controlled trial [ClinicalTrials.gov: NCT03836625]. The trial assessed the feasibility, acceptability and initial efficacy of using CareConekta, a smartphone app designed to facilitate engagement in HIV care by enabling users to locate nearby ART facilities in times of mobility. The trial recruited pregnant WLHIV from the Gugulethu Midwife Obstetric Unit (MOU), a public antenatal care (ANC) clinic based in the community of Gugulethu in Cape Town, South Africa. This is a high HIV burden setting, with an estimated antenatal HIV prevalence of 30%.^24^ Participants were enrolled in the third trimester of pregnancy and followed through six-months postpartum. In this analysis, using data from the screening/enrolment visit, we describe smartphone ownership patterns and use among screened eligible and ineligible WLHIV.

### Study recruitment and eligibility

All pregnant WLHIV attending the Gugulethu MOU interested in learning about the study were screened for eligibility. Eligibility criteria included being at least 18 years old, pregnant (≥28 weeks gestation), living with HIV, able to speak and understand isiXhosa (predominant local language) and/or English, currently own a smartphone that met certain technical requirements, willing to opt-in to app installation and mobility tracking, and able to demonstrate basic smartphone literacy. For the purposes of this study, a smartphone was defined as a mobile phone with a touchscreen interface and internet and Global Positioning System (GPS) capabilities. In order to enrol, eligible women needed to have their phone available with them at the time of enrolment. Specific technical requirements for enrolment that were assessed at screening included whether the phone had the Android operating system version 5.0 or later, connection to one of the four major cell phone service providers (Vodacom, Cell-C, Telkom or MTN), able to demonstrate use of GPS by opening a map app and finding the current location and requiring charging less than twice per day (by self-report).

### Measures

An eligibility checklist form was completed to determine eligibility [**Supplemental Materials**]. On this form, those found ineligible, or who were eligible but declined participation, were asked to answer a short list of demographic questions following verbal consent. The eligibility checklist was designed to administer all questions on the form, even after the earliest indication of being ineligible. However, administration of this was not always consistent; thus, 18% of participants did not complete all the screening questions after they were deemed ineligible. Consequently, there is variation in sample size between each variable on the eligibility checklist.

Eligible women willing to participate in the trial completed written informed consent and were enrolled in the CareConekta study. Enrolment was followed by an interviewer-administered questionnaire, which collected similar demographic information and asked about smartphone ownership and usage. All study measures were available in English and isiXhosa.

First, to describe the mHealth landscape among the full population of pregnant WLHIV attending care at the MOU, we compared the smartphone ownership characteristics of those who were enrolled in the CareConekta trial (i.e., met all the smartphone-related eligibility criteria) and those who were not enrolled. Those who were not enrolled included all those who did not meet the CareConekta eligibility criteria and those who were eligible but did not enrol on the day of screening, either because they were not interested, were referred to another facility or were not available to enrol on the day of screening.

Next, to explore differences in sociodemographic characteristics among those enrolled versus not enrolled, we present data grouped by enrolment status, and further, by smartphone ownership status: those who owned smartphones versus those who did not (including those who did not own a mobile phone at all).

Lastly, to explore smartphone usage and preferences among women enrolled in the CareConekta trial, we asked about phone ownership and sharing patterns, common phone uses and popular apps, cell phone and cell number turnover and common reasons for the phone being ‘off’ for extended periods of time.

### Data analysis

Data were analysed using STATA 14.2 (Stata Corporation, College Station, Texas). Descriptive statistics were used to describe the baseline characteristics of all screened women as well as smartphone ownership and usage patterns, presented as frequencies and proportions.

Bivariate analyses were conducted using the Wilcoxon Rank-Sum test for the numerical variables and the Chi-squared or Fishers exact tests for categorical variables. For all analyses, a threshold p-value of 0.05 and 95% confidence intervals (CI) were used to determine statistical significance.

## Results

A total of 661 WLHIV were screened for enrolment into the trial between December 2019 and February 2021. For this analysis, we excluded the screening data of the first 22 women to be screened due to incomplete data. Therefore, a total of 639 women were included in this analysis. Overall, 200 women were enrolled in the CareConekta trial. However, women for whom app installation failed at enrolment (n=6) and those who no longer met the eligibility criteria within 14 days of enrolment (n=1) were withdrawn from the trial and are excluded from the analysis of enrolled participants (n=193).

### Smartphone ownership patterns and determinants of eligibility for the CareConekta trial

**Table 1** details the determinants of eligibility for the CareConekta trial. Out of 639 women included in this analysis; 259 (40.5%) met eligibility criteria and 380 did not. Among those eligible, 59 were not enrolled (because they were either not interested, needed time to think about enrolling or promised to return on another day for enrolment).

**Table 1.** Determinants of eligibility and smartphone ownership patterns among 639 women who were screened for the CareConekta study.

| **Variable** | **Total N=639** | **\*Ineligible N=380** |
| --- | --- | --- |
| **Third trimester (n=638)** |  |  |
| Yes | 412 [64.6] | 153 [40.4] |
| No | 226 [35.4] | 226 [59.6] |
| **English/IsiXhosa Understanding (n=637)** |  |  |
| Yes | 619 [97.2] | 360 [95.2] |
| No | 18 [2.8] | 18 [4.8] |
| **Age: Median [IQR]** | 31 [27-35] | 31 [26-35] |
| **Mobile phone ownership (n=637)** |  |  |
| Yes | 582 [91.4] | 323 [85.4] |
| No | 55 [8.6] | 55 [14.6] |
| **Smartphone ownership (n=580)** |  |  |
| Yes | 503 [86.7] | 244 [76.0] |
| No | 77 [13.3] | 77 [24.0] |
| **Android operating system version 5.0 or later (n= 458)** |  |  |
| Yes | 421 [91.9] | 162 [81.4] |
| No | 37 [8.1] | 37 [18.6] |
| **Service provider (Vodacom, Cell-C, Telkom or MTN) (n=506)** |  |  |
| Yes | 504 [99.6] | 245 [99.2] |
| No | 2 [0.4] | 2 [0.8] |
| **GPS-enabled phone (n=506)** |  |  |
| Yes | 497 [98.2] | 238 [96.4] |
| No | 9 [1.8] | 9 [3.6] |
| **Battery strength (n=506)** |  |  |
| Charged < twice a day | 485 [95.9] | 226 [91.5] |
| Charged > twice a day | 21 [4.1] | 21 [8.5] |
| **Phone with participant at screening (n=507)** |  |  |
| Yes | 438 [86.4] | 179 [72.2] |
| No | 69 [13.6] | 69 [27.8] |

\*Reasons for ineligibility do not sum up to 100% because some respondents fit into more than one category of ineligibility.

Among screened women (n=639), age ranged from 16-45 years, with a median age of 31 years (interquartile range [IQR]: 27-35 years). Most women screened (91.4%) owned mobile phones and 86.7% of those owned smartphones. Among those who owned smartphones and responded to the relevant ‘smartphone feature’ questions (n=458), 91.9% owned phones with an Android operating system version 5.0 or above, all used one of the four most common local network service providers, 98.2% used phones that had a GPS feature, 95.9% charged their phones less than twice a day, and 86.4% had their phones with them at the time of screening.

Among those ineligible (n=380), being outside the gestational age window for the trial was the most common reason for ineligibility (59.6%), followed by not having the smartphone with them at the time of screening (27.8%), and lastly the phone not being a smartphone (24.0%).

### Socio-demographic characteristics of women who own versus those who do own not smartphones

A total of 550 women had complete sociodemographic data available, including 193 women enrolled in the trial and 357 women not enrolled who answered the questions at screening. **Table 2** shows women’s socio-demographic characteristics, stratified by smartphone ownership status.

**Table 2.**
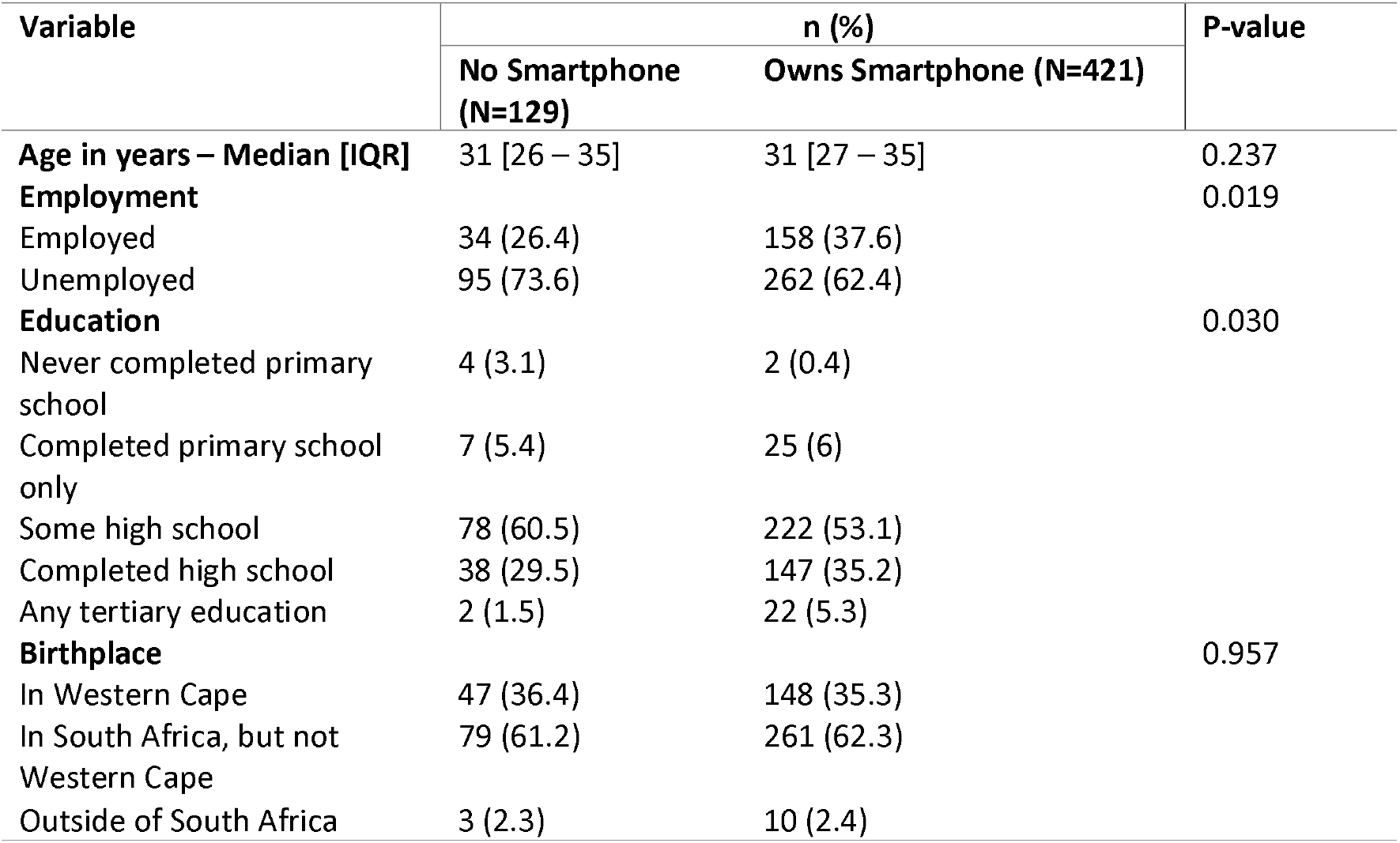
Sociodemographic characteristics among women who were screened for the CareConekta study, by enrolment status and smartphone ownership.

Overall, 76.5% (n=421) of women owned a smartphone. Median age was 31 years in both groups (p=0.237). Unemployment was high overall, but significantly higher among those who did not own a smartphone (73.6%) compared to those who did (62.4%), p=0.019. Most women had completed some high school education, with those who owned smartphones more likely to have reached higher levels of education compared to those who do not own smartphones (35.2% versus 29.5% completed high school, p=0.030). Two-thirds of participants were born outside of the Western Cape province (where the study clinic was located), with no difference by smartphone ownership (p=0.957).

### Smartphone ownership and use patterns among women enrolled in the CareConekta study

**Table 3** describes smartphone ownership and use patterns among the 193 enrolled participants. Most participants (98.5%) owned their own smartphones. A minority of participants (13.5%) shared their smartphone with someone; of these, phone sharing was most common with a romantic partner (71.4%), followed by a family member (28.6%). In these cases, nearly all participants (96%) reported having the phone with them most of the day. Median duration of ownership of the particular smartphone used at enrolment was 12 months [IQR: 5-24], median duration of use of the phone number at enrolment was 25 months [IQR: 12-60], and median number of cell phone numbers participants owned in the two years prior to enrolment was two [IQR: 1-2].

**Table 3.** Smartphone use patterns among women who were enrolled in the CareConekta study.

| **Variable** | **n (%)** |
| --- | --- |
| **Phone ownership (n=193)** |  |
| Owned by participant | 190 (98.5) |
| Owned by intimate partner | 2 (0.5) |
| Owned by family member | 1 (1.0) |
| **Phone sharing (n=193)** |  |
| Phone not shared | 167 (86.5) |
| Phone shared | 26 (13.5) |
| **Person phone is shared with (n=26, more than one response allowed):** |  |
| Romantic partner | 20 (71.4) |
| Family member | 8 (28.6) |
| **Person who has phone most of the day if shared (n=26)** |  |
| Participant | 25 (96.2) |
| Someone else | 1 (3.8) |
| **Duration of phone ownership in months - Median [IQR]** | 12 [5-24] |
| **Duration of cell number use - Median [IQR]** | 25 [12-60] |
| **Number of cell phone numbers in past two years** | 2 [1-2] |
| **Phone use (n=193)☐** |  |
| Receive phone calls | 192 (99.5) |
| Receive SMSs | 191 (99.0) |
| Send SMSs | 191 (99.0) |
| Make phone calls | 190 (98.5) |
| Send 'Please call me' | 184 (95.3) |
| **Instant messaging (i.e., WhatsApp, Facebook messenger)☐** | 182 (94.3) |
| **Photos** | 180 (93.3) |
| **Internet** | 146 (75.7) |
| **Email** | 91 (47.2) |
| **GPS (Maps)** | 106 (54.9) |
| **Other** | 74 (38.3) |
| **Favourite Apps (n=193)☐** |  |
| WhatsApp | 181 (93.8) |
| Facebook | 112 (58.0) |
| Instagram | 10 (5.2) |
| Twitter | 5 (2.6) |
| Other | 35 (18.1) |

☐ More than one response was allowed, thus, numbers sum to over 100%

The most common smartphone uses reported were receiving (99.5%) and making (98.5%) phone calls, receiving and sending SMSs (99.0%), sending ‘please call me’ requests (95.3%) and instant messaging apps, such as WhatsApp and Facebook messenger (94.4%). A ‘please call me’ request is a free standardized text message (provided by mobile network companies) that allow one user to alert another to give them a call. Conversely, the least used features were GPS (54.9%) and email (47.2%). When asked about their favourite apps (and allowed to tick all that apply), WhatsApp was most popular by far (93.8%), ranking well above Facebook (58.0%), Instagram (5.2%) and Twitter (2.6%).

## Discussion

As mHealth studies continue to grow worldwide, including in low-resource settings, it is important to examine the landscape of mobile phone and smartphone users in a routine public health setting. In our study in South Africa, mobile phone ownership was found to be very common, with nearly all (91.7%) of WLHIV surveyed owning mobile phones and 86.7% of WLHIV owning smartphones. This is consistent with existing literature on smartphone ownership in Sub-Saharan Africa, including South Africa;^25^ however our findings represent a sample of WHLIV attending antenatal care in the public sector, so are of specific interest to those developing mHealth interventions for similar populations.

In our study, some socio-demographic characteristics differed by smartphone ownership. Consistent with a national survey, we found that smartphone owners had higher education and employment than non-owners.^25^ We did not find significant differences in age nor by place of birth. Researchers considering mHealth studies in similar settings should be aware that they may miss potentially vulnerable populations with lower education and employment levels by requiring smartphones.

Although having the potential to be accessible and cost-effective, mHealth interventions are not without their challenges. Earlier research showed that some of the biggest challenges of mHealth interventions in Sub-Saharan Africa include inconsistent electricity and network coverages, phone sharing, poor phone quality and battery life, as well as high mobile phone and phone number turnover.^26^ In our findings, most of the smartphones met the most basic technical requirements for installation of a mobile app. As expected, nearly all (91.9%) of smartphones used the Android operating system, and 95.9% were reported to require one or fewer battery charges per day, which is an encouraging finding for future mHealth interventions in peri-urban settings. Nearly all (98.2%) smartphones included a GPS feature that is conducive for location-based mHealth interventions, such as CareConekta.

We also found that phone sharing was not common in this setting. Furthermore, in nearly all the cases where phone sharing occurred, the phone was being shared with a romantic partner or family member and the phone was with the owner for most of the day. These findings are consistent with literature on phone sharing patterns in similar populations.^27,28^ Limited phone sharing is encouraging for mHealth interventions because it minimizes periods of loss of engagement with the intervention, as well minimizing opportunities for privacy and confidentiality loss. This is particularly important for interventions centred around HIV management, especially given the persistence and complexities of HIV-related stigma in countries like South Africa; a common barrier to HIV adherence.^29^ Although phone sharing was found to be uncommon, mHealth interventions should remain sensitive to issues of user privacy and confidentially.

We found variability in the evidence of smartphone and phone number turnover in this population, but overall turnover of device and phone numbers was moderate. Our findings on smartphone turnover are consistent with those of a 2015 study on smartphone and phone number turnover among pregnant WLHIV in South Africa.^28^ However, compared to the same study, which reported that most participants had used their current phone number for three years and some as long as thirteen years, we found slightly higher turnover.^28^ Furthermore, the earlier study found that most participants used the same phone number in the past two years, we found that most of our participants had used two numbers in the past two years^28^. Therefore, phone and phone number turnover needs to be considered in future mHealth interventions in similar settings. This includes designing mHealth applications that are easy and cheap to re-install, as well as easy to re-register and flexibility to access pre-existing profiles.

Participants reported that making and receiving calls, as well as sending and receiving text messages, were their most used features. This is consistent with an earlier study in this setting that found that text messaging was the most commonly used feature.^25^ Texting can be leveraged for the most basic mHealth interventions, i.e. health education or adherence reminder interventions, that require minimal-to-no direct reciprocal engagement from the individual. Furthermore, this feature is commonly found in any mobile phone – thus having the potential to reach both smartphone and non-smartphone users.

GPS and email usage were the least common features used. Low email usage is contrary to the qualitative study by Mogoba et al. in the same setting that showed email to be among the top smartphone uses but consistent with quantitative findings that found email usage to be the least common.^27,28^ Email usage or the ownership of an email address, at the very least, is important to consider for mobile application mHealth interventions because installation from platforms such as Google Play store requires a Google-authenticated email address.

We also found that the existing messaging platform WhatsApp is incredibly popular in this setting, with Facebook coming in a distant second. This finding is consistent with literature in similar settings.^27,28^ Future mHealth interventions may wish to leverage the popularity of these already existing apps since people are already familiar with installing and using them, and are already contributing financially to use these apps.

These findings should be viewed with an understanding of the strengths and limitations of our research. The CareConekta study specifically enrolled pregnant women who were living with HIV and attending ANC; it is possible that the individuals seeking care other than ANC differ in smartphone ownership and use patterns. The CareConekta study, however, approached every WLHIV who was attending ANC and thus this analysis is likely to be representative of this specific population of women. Missing data on eligibility checklist forms at the beginning stages of the CareConekta study resulted in the exclusion of some data for analysis in this research. Lastly, a strength of this research is that we explored the technical specifications of these smartphones which is not common in the literature and provides more data to support future mHealth intervention design.

In conclusion, we found that smartphone ownership is very common in this low-resource, peri-urban setting. Common mHealth challenges such phone sharing, poor battery life to support mHealth applications, and smartphone and phone number turnover were either uncommon or moderate. These results are encouraging to the development and roll-out of mHealth interventions in similar settings. Our findings on the popularity of existing platforms, particularly WhatsApp, are also encouraging and should be leveraged for mHealth interventions.

## Data Availability

All data produced in the present study are available upon reasonable request to the authors.

## Acknowledgements

The authors are most grateful to the study participants, without whom this work would not be possible. We also wish to acknowledge the Western Cape Department of Health and their provincial data centre. This work was supported by the US National Institute of Mental Health (R34 MH118028). T.K.P. is supported by a CIPHER grant from the International AIDS Society and the Fogarty International Center and National Institute of Mental Health of the National Institutes of Health under Award Number K43TW011943. The content is solely the responsibility of the authors and does not necessarily represent the official views of the National Institutes of Health or other parties.

