## Supplemental Materials for "Smartphone ownership and use among pregnant women with HIV in South Africa"

Eligibility checklist (English version; also available in isiXhosa)

We are conducting a study to assess a new smartphone application (“app”) called CareConekta. We designed CareConekta to help postpartum women living with HIV link to HIV care services if they travel away from their home area. We want to assess the feasibility and acceptability of using this new smartphone app and also see if it helps to link women to care.

If you think you might be interested, I will ask you a few demographic questions, and then check that you are eligible for the study and then we will give you all the information you need to decide if you would like to take part. Participation is voluntary and you can decide not to take part at any time without any penalty.

|  | We will be recruiting 200 women to try out this new application for about 9 months. The app will be loaded on your smartphone for free – do you think you might be interested? | ___ Yes  ___ No  →  continue to demographic questions |
| --- | --- | --- |
|  | Date screening completed | __ __ \| __ __ __ \| __ __ __ __ |
| *Eligibility screening* | | |
|  | How old are you?  [Confirm using clinic folder or ID document] | __________ years  If not yet 18 years, NOT ELIGIBLE |
| 2. | Do you own a mobile phone? | 0=No NOT ELIGIBLE  1=Yes CONTINUE |
| 6. | Confirm by looking at her phone | 0=not confirmed  1=confirmed  → CONTINUE |
| 7. | Are you ≥28 weeks pregnant?  [Confirm in maternity case record] | 0=No → NOT ELIGIBLE – invite to review again at next clinic visit  1=Yes → CONTINUE |
|  | Are you HIV-positive?  [Confirm in maternity case record] | 0=No →  NOT ELIGIBLE  1=Yes →  CONTINUE |
|  | What is your preferred language? | 1=isiXhosa  2=English  3=Other, specify: _______________________ |
|  | Are you able to read basic isiXhosa or English?   - Confirm by asking the enrolee to read the following sentence in English or isiXhosa:   “I can look up a new clinic on the phone.” | 0=No → NOT ELIGIBLE  1=Yes → CONTINUE |
|  | Based on the above questions is she eligible for study? | 0=No → Continue to demographic questions  1=Yes → refer to study team for informed consent |

Demographics Questionnaire (English version; also available in isiXhosa)

| **Demographics of women who are ineligible or declined** | | |
| --- | --- | --- |
| Even though you are not eligible for the study or prefer not to take part, I would like to ask you a few basic demographic questions. These answers will be anonymous and will be used to describe the group of women who did not participate in the study.  Is that OK with you? | | ___ Yes → Continue  ___ No →Thank you for your time! |
| ***Demographic information*** | | |
| 1. | How old are you? | __________ years |
| 2. | Are you currently employed? | ___ Yes  ___ No |
| 3. | What is the highest level of schooling that you have completed? | ___ Never completed primary school (did not finish grade 7/standard 5  ___Completed primary school only (finished grade 7/standard 5)  ___Some high school (grade 8-11/standard 6-9)  ___Completed high school (matric/grade 12/standard 10)  ___Any tertiary education |
| 4. | Where were you born? | ___In Western Cape  ___In South Africa, but not Western Cape  *Specify province:*  ________________________  Not in South Africa  *Specify country:* ____________________________ |
| 5. | Write phone make/model and IMEI number | Make:___________________________  Model:__________________________  IMEI:____________________________ |
| 6. | Why would you not like to participate? [only for those who did not wish to participate] |  |
